# Profiling Zero-Dose Measles-Rubella Children in Zambia: Insights from the 2024 Post-Campaign Coverage Survey

**DOI:** 10.1101/2025.09.16.25335955

**Authors:** Moses Mwale, Guissimon Phiri, Francis Dien Mwansa, Peter J. Chipimo, Penelope Masumbu, Kennedy Matanda, Princess Kayeye, Kelvin Mwangilwa, Chola Nakazwe, Harriet Namukoko, Simon Mutembo, Freddie Masaninga, Jacob Sakala, Peter Clement Lugala

## Abstract

**Background:** Measles & Rubella Zero-dose children (unvaccinated for measles– rubella) cluster in underserved communities and sustain measles transmission. We estimated MR zero-dose prevalence after Zambia’s 2024 MR Supplementary Immunisation Activity (SIA) and identified associated risk factors and barriers.

**Methods:** A coverage survey (two-stage stratified cluster design) across all 10 provinces, 27 December 2024–16 January 2025, involved interviewing caregivers of children aged 9–59 months; vaccination status was verified by card (11.7%) or recall (88.3%). Data were analyzed using survey-weighted methods and logistic regression, adjusting for stratification, clustering, and sampling weights.

**Results:** Among 8,634 children, MR zero-dose prevalence was 11.97% (95% CI: 11.03–12.91), highest in Central (19.15%) and Western (17.71%), lowest in Copperbelt (6.69%). Urban residence reduced odds by 24% vs. rural (aOR 0.76, 95% CI: 0.63–0.92). Risks rose with age (>36 months: aOR 1.60, 95% CI: 1.27– 2.00), maternal absence (aOR 1.74, 95% CI: 1.33–2.27), or death (aOR 2.40, 95% CI: 1.23–4.68). Most zero-dose children (88.75%) lacked other vaccines, indicating systemic gaps. Key barriers included unawareness (42.58%) and travel time (>2 hours: aOR 3.20, 95% CI: 1.43–7.16).

**Conclusions:** Nearly one in eight Zambian children remained MR zero-dose post-2024 SIA, concentrated in rural, high-prevalence areas, older children, and motherless households. Priorities include health worker-led awareness campaigns, mobile services to cut travel time, and integrated SIA-RI strategies (microplanning, tracing, catch-up) to address systemic gaps, supporting global measles elimination under IA2030.

## Introduction

Measles remains one of the most contagious vaccine-preventable diseases and continues to cause substantial morbidity and mortality worldwide. In 2022 alone, measles caused approximately 128,000 deaths globally, the vast majority among unvaccinated children in low-and middle-income countries (LMICs) (1). Despite progress, 14.3 million children globally were classified as “zero-dose” in 2024—that is, they had not received any dose of a routine vaccine. This group remains a major driver of measles transmission through clustering in under-immunised communities (2,3). Outbreaks in both LMICs and high-income countries underscore the continued threat posed by declining or stagnant immunisation coverage (4).

Zambia is no exception to these challenges. Between 2021 and 2024, the number of measles zero-dose children in Zambia rose from approximately 39,000 to 41,000, ranking the country among the top 12 in the Eastern and Southern Africa Region (ESAR) (5). Nationally, measles elimination requires sustained ≥95% coverage with two doses of the measles-rubella (MR) vaccine, as outlined in the World Health Organization’s (WHO) *Immunization Agenda 2030 (IA2030)* (6). However, routine immunisation coverage has stagnated, and inequities persist, especially in rural and socioeconomically disadvantaged communities. These gaps threaten Zambia’s ability to achieve measles elimination goals and mirror global concerns about stagnating immunisation progress (7,4).

Several studies have identified risk factors for zero-dose status, including rural residence, maternal absence or orphanhood, caregiver education level, and distance to health facilities (8,9,10). However, many analyses rely on secondary datasets, lack disaggregated campaign-specific information, or fail to capture behavioural and social determinants of vaccine uptake. This limits the ability of policymakers to design context-specific interventions to close immunisation gaps. Locally generated evidence is especially critical as both Gavi’s equity agenda and IA2030 prioritise reaching zero-dose children as a global public health imperative (6,11).

In Zambia, the 2024 Measles–Rubella Supplementary Immunisation Activity (SIA) was reported to have reached 97% coverage administratively, vaccinating 165,000 zero-dose children through targeted microplanning, community engagement, and digital monitoring (12). Yet, administrative coverage often overestimates true performance. Independent evaluation is necessary to validate coverage, quantify remaining zero-dose prevalence, and identify risk factors limiting access and uptake. To address these gaps, a Post-Campaign Coverage Survey (PCCS) was conducted following Zambia’s 2024 MR SIA. The survey was designed to provide an independent estimate of vaccination coverage among children aged 9–59 months, offering a validated measure beyond routine administrative reports. It also aimed to quantify the prevalence of zero-dose and under-immunised children, assess their overlap with routine immunisation performance, and evaluate the proportion reached during the campaign. In addition, the PCCS examined socio-demographic, geographic, and awareness-related factors associated with non-vaccination, generating insights into barriers to uptake and informing strategies to reduce missed opportunities for immunisation. By generating robust and nationally representative evidence, this study provides critical insights for strengthening measles control in Zambia and contributes to the global evidence base on strategies to reduce zero-dose children in LMICs.

## Methods

### Study Design

The 2024 MR Post-Campaign Coverage Survey (PCCS) was a cross-sectional household survey to evaluate the September 2024 Measles–Rubella (MR) Supplementary Immunisation Activity (SIA) in Zambia. The survey followed the WHO Vaccination Coverage Cluster Survey Reference Manual (13) and was implemented between 27 December 2024 and 16 January 2025 across all 10 provinces. Zambia has an estimated population of 19,610,769 (14), administratively divided into provinces, districts, constituencies, and wards. Within wards, enumeration areas (EAs) mapped during the 2022 Census of Population and Housing served as the primary sampling units.

### Target Population

The survey targeted children aged 9–59 months at the time of the September 2024 SIA. Eligible children were those born between 23 October 2019 and 23 December 2023. Household eligibility required at least one child under the age of seven years residing in the household.

### Sampling Frame and Sample Design

The sampling frame was derived from the 2022 Census of Population and Housing. A two-stage stratified cluster sampling design was employed. In the first stage, 370 EAs (clusters) were selected using probability proportional to size (PPS), stratified by province and by urban/rural residence. In the second stage, systematic random sampling was used to select 25 households with at least one eligible child from each EA, after a complete household listing exercise. This yielded a planned sample of 9,250 households, providing adequate precision for national, provincial, and urban– rural level estimates.

### Sample Weights

Sampling weights were calculated as the inverse of the product of the first-stage and second-stage selection probabilities. Adjustments were applied for non-response at the household level. All analyses incorporated sampling weights to generate population-representative estimates.

### Data Collection Tools

A structured household questionnaire, adapted from the WHO PCCS standard tool, was programmed into the CSPro application for Computer-Assisted Personal Interviewing (CAPI). The instrument included modules on household demographics, caregiver education and occupation, child vaccination history (card verification or caregiver recall), knowledge and awareness of the SIA, sources of information, and barriers to vaccination uptake. Behavioural and social drivers of vaccination were assessed using items aligned with the WHO BeSD (Behavioural and Social Drivers of Vaccination) framework.

### Training and Fieldwork

A total of 164 field staff (131 enumerators and 33 supervisors) underwent a six-day centralised training facilitated by the Zambia Statistics Agency and Ministry of Health. Training included survey objectives, questionnaire content, CAPI procedures, household listing, interviewing techniques, ethical considerations, and practical field exercises. Only participants who passed competency assessments were deployed. Field teams of 4–6 members conducted household listing and interviews under close supervision. Geographic Information Systems (GIS) staff verified EA coverage before household selection. Data were collected electronically using tablets, with secure daily uploads to a central server at the Zambia Statistics Agency.

### Data Quality Assurance

Supervisors conducted daily field checks and verified completed interviews. Automated data consistency checks were embedded in the CAPI system. Data files were reviewed centrally for completeness, and errors were corrected before final cleaning. Pictures of vaccination cards were taken, where available, to validate caregiver reports.

### Response Rates

Of the 9,211 sampled households, 9,018 were successfully interviewed, yielding a household response rate of 97.8% (98.1% urban; 97.6% rural). A total of 8,634 eligible children were surveyed, representing 2.17 million children nationally when weighted.

### Statistical Analysis

Survey-weighted analyses were performed using the *survey* package in R (version 4.3.2). Proportions and 95% confidence intervals (CIs) were estimated, accounting for clustering and stratification. Logistic regression models were used to identify predictors of zero-dose status, including demographic, socio-economic, and access-related factors. Statistical significance was set at p < 0.05. Missing data were retained in descriptive summaries but excluded from regression models.

### Ethical Considerations

The PCCS was implemented under the authority of the Zambia Ministry of Health as a public health surveillance activity and was not classified as human subjects research. Nevertheless, ethical principles were observed. Informed verbal consent was obtained from all caregivers prior to interviews. All data were anonymised, and personal identifiers were not retained in analytic datasets.

## Results

### Zero-Dose Prevalence

Nationally, 11.97% (95% CI: 11.03–12.91%) of children were MR zero-dose, of whom 88.75% had not received DPT1 or other antigens, indicating systemic immunization gaps (Supplementary Table S2). Prevalence was highest in Central (19.15%) and Western (17.71%) provinces, and lowest in Copperbelt (6.69%) (Figure 1). Rural prevalence (13.02%) exceeded urban (10.17%), with an adjusted odds ratio (OR) for urban residence of 0.76 (95% CI: 0.63–0.92), indicating higher odds of zero-dose status in rural settings (Supplementary Table S1). Children in large households (≥9 members) had a higher MR zero-dose prevalence (14.18%; 95% CI: 11.63–16.73%) compared to those in households with 1–4 members (12.29%; 95% CI: 10.72–13.86%) or 5–8 members (11.17%; 95% CI: 9.88–12.46%), though this association was not statistically significant (Supplementary Table S1). Stratified analysis showed similar patterns in both rural (≥9: 14.41%) and urban settings (≥9: 13.41%), confirming that the household size effect was consistent but not significant across residence types.

**Table 1.** Key findings on MR zero-dose prevalence and adjusted associations, Zambia PCCS 2024 (n = 8,634)

| Variable (comparison vs reference) | Zero-dose prevalence (%) | 95% CI (%) | Adjusted OR (95% CI) |
| --- | --- | --- | --- |
| National prevalence | 11.97 | 11.03–12.91 | — |
| Province – Central | 19.15 | 15.84–22.46 | — |
| Province – Copperbelt | 6.69 | 4.67–8.71 | — |
| Urban (vs Rural) | 10.17 | 9.29–11.05 | 0.76 (0.63–0.92) |
| Age >36 months (vs 0–12 months) | 14.99 | 13.25–16.73 | 1.60 (1.27–2.00) |
| Mother absent (vs present) | 19.73 | 15.85–23.61 | 1.74 (1.33–2.27) |
| Mother deceased (vs present) | 24.17 | 12.60–35.74 | 2.40 (1.23–4.68) |
| Travel time >2 hours (vs <15 minutes) | 25.71 | 10.94–40.48 | 3.20 (1.43–7.16) |
| Campaign awareness – Not heard (vs Heard) | 18.50 | 16.17–20.83 | 2.00 (1.65–2.42) |

**Notes:**
1. Prevalence values are zero-dose prevalence within each subgroup (survey-weighted, with 95% CIs). 2. Adjusted ORs are from survey-weighted logistic regression accounting for stratification, clustering and sampling weights; reference categories shown in brackets. 3. Reasons for non-receipt (e.g., “Lack of awareness 42.58%”) describe the distribution of reported reasons among zero-dose children and are not directly comparable to subgroup prevalence; report these in a separate table/figure. 4. Systemic gap marker: *Among zero-dose children, 88.75% had received no other routine vaccines.* If desired, add a one-line footnote: “Receipt of other routine vaccines (vs none) was associated with lower odds of MR zero-dose (aOR 0.26, 95% CI 0.08–0.91).”

**Figure 1.**
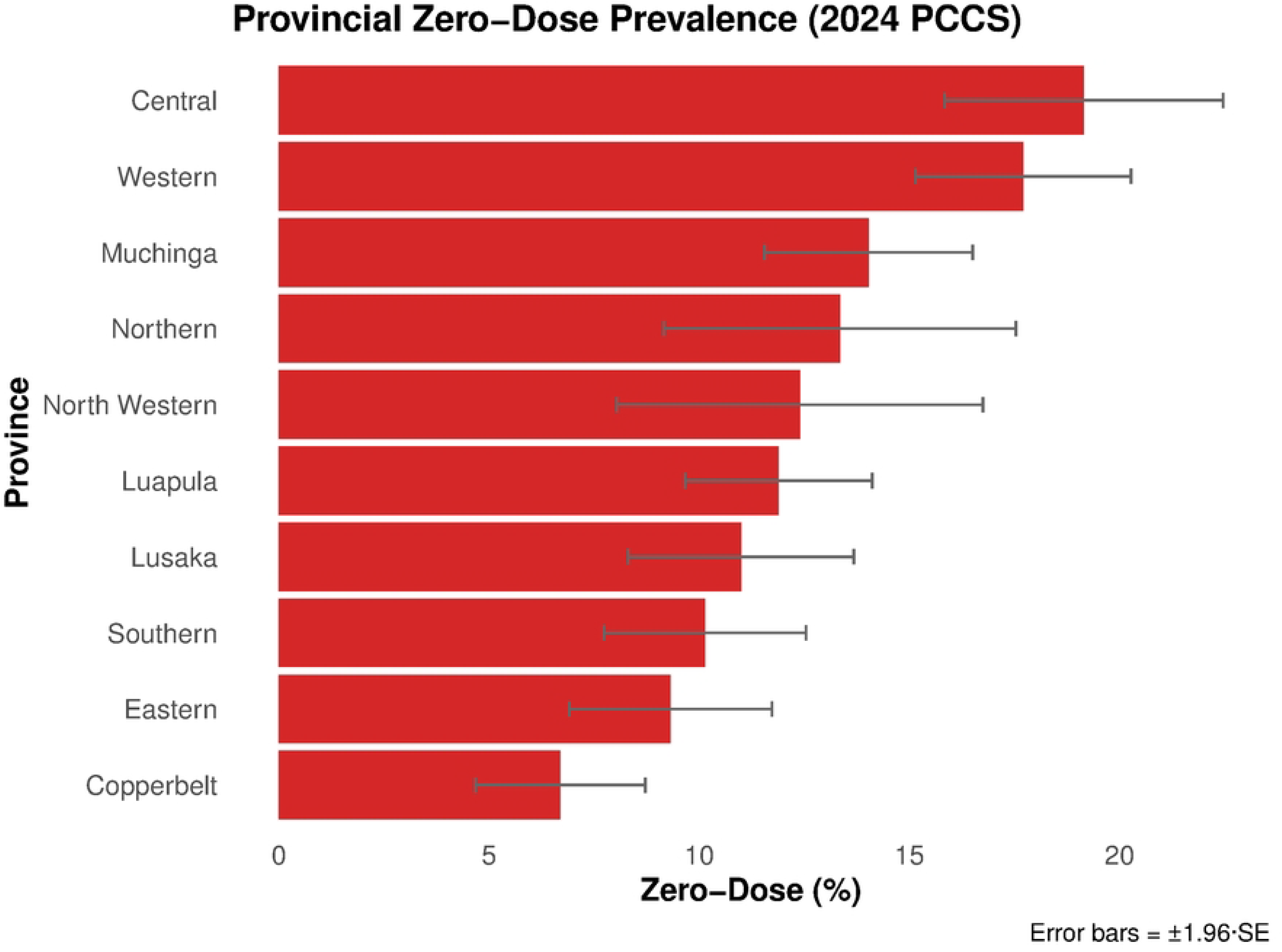
Weighted Zero-Dose Measles-Rubella Prevalence by Province, Zambia, 2024 PCCS. Exact values are in Supplementary Table S1.

### Demographic Risk Factors

Measles-rubella (MR) zero-dose prevalence increased with age, peaking at 14.99% among children >36 months (OR = 1.60; 95% CI: 1.27–2.00) (Figure 2). No significant sex differences were observed between males (11.90%; 95% CI: 10.92– 12.88%) and females (12.04%; 95% CI: 11.04–13.04%), with an adjusted odds ratio of 1.05 (95% CI: 0.89–1.23; p = 0.533) (Supplementary Table S1). Children whose mothers were absent (19.73%; OR = 1.74, 95% CI: 1.33–2.27) or deceased (24.17%; OR = 2.40, 95% CI: 1.23–4.68) had increased odds of being MR zero-dose (Supplementary Table S1). A small group with “unknown” maternal status showed zero vaccination (OR = 0.00), but this reflects unstable estimates due to very small sample size. Guardian education (e.g., Primary: 11.33%; 95% CI: 9.82–12.84%) and religion (e.g., Protestant: 12.50%; 95% CI: 11.44–13.56%) showed no statistically significant associations (Supplementary Table S2). Similarly, socioeconomic status (SES), derived from occupation, did not show a consistent pattern across low, medium, and high categories, and associations were not statistically significant. Muslim prevalence was unreliable (0.00%) due to small sample size (Supplementary Table S2).

**Figure 2.**
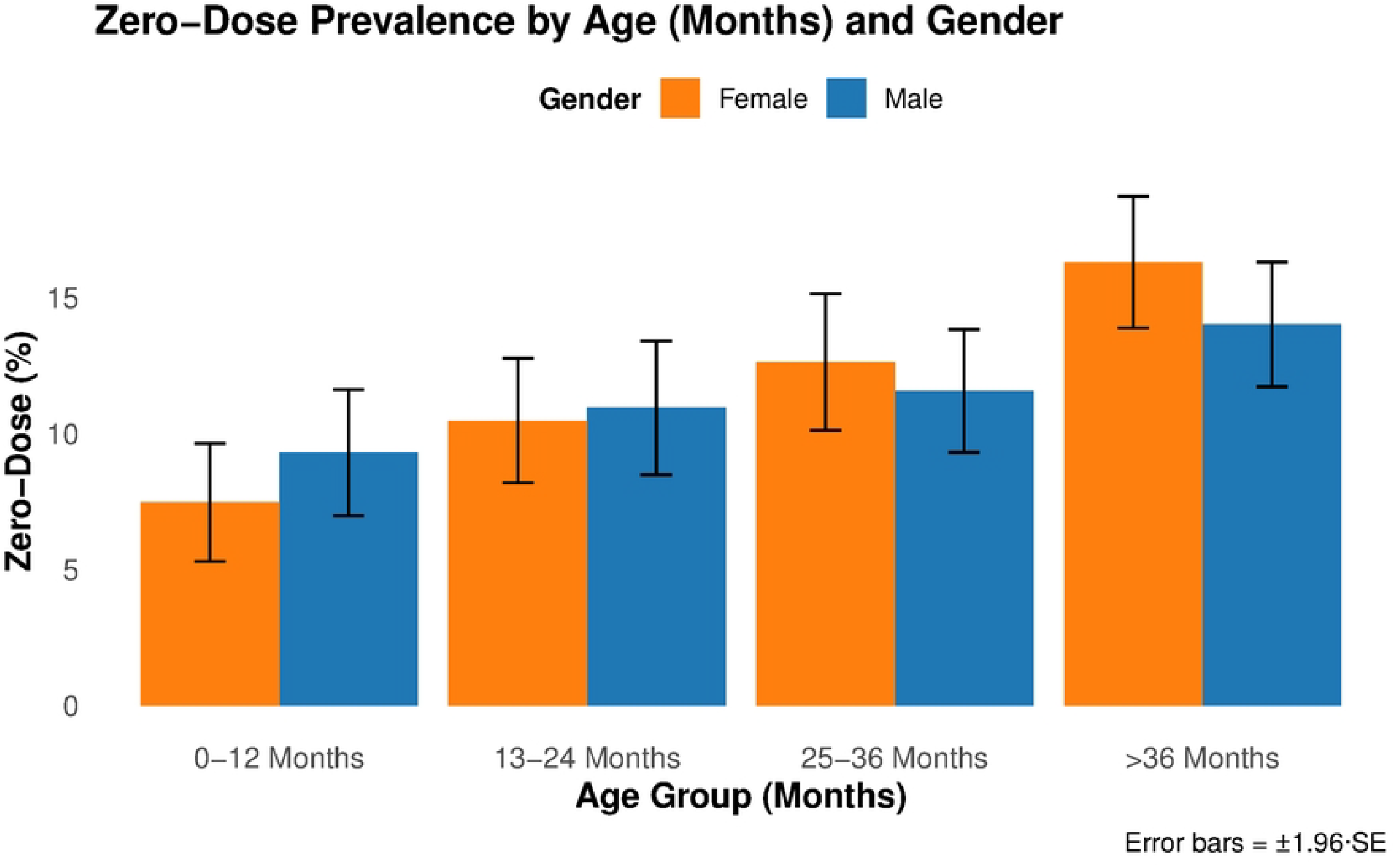
Weighted Percentage of MR Zero-Dose Children by Age and Gender, Zambia, 2024 PCCS.

### Systemic Vaccination Gaps

Among MR zero-dose children, 88.75% had not received any other basic vaccines, indicating broader immunisation failure rather than isolated MR campaign gaps. Those who received other vaccines had significantly lower odds of being MR zero-dose (OR = 0.26; 95% CI: 0.08–0.91).

### Barriers to Vaccination

Lack of awareness of the campaign (42.58%) was the most cited reason for non-vaccination, followed by being too busy (19.02%) and inconvenient timing (7.69%) (Figure 3). Travel time of 1–2 hours (OR = 1.55; 95% CI: 1.20–2.00) and over 2 hours (OR = 3.20; 95% CI: 1.43–7.16) were associated with significantly higher odds of being MR zero-dose (Supplementary Table S2). The regression model also included travel means (mode of transport to the facility), but no significant associations were observed. CIs for >2 hours were wide due to small sample sizes.

**Figure 3.**
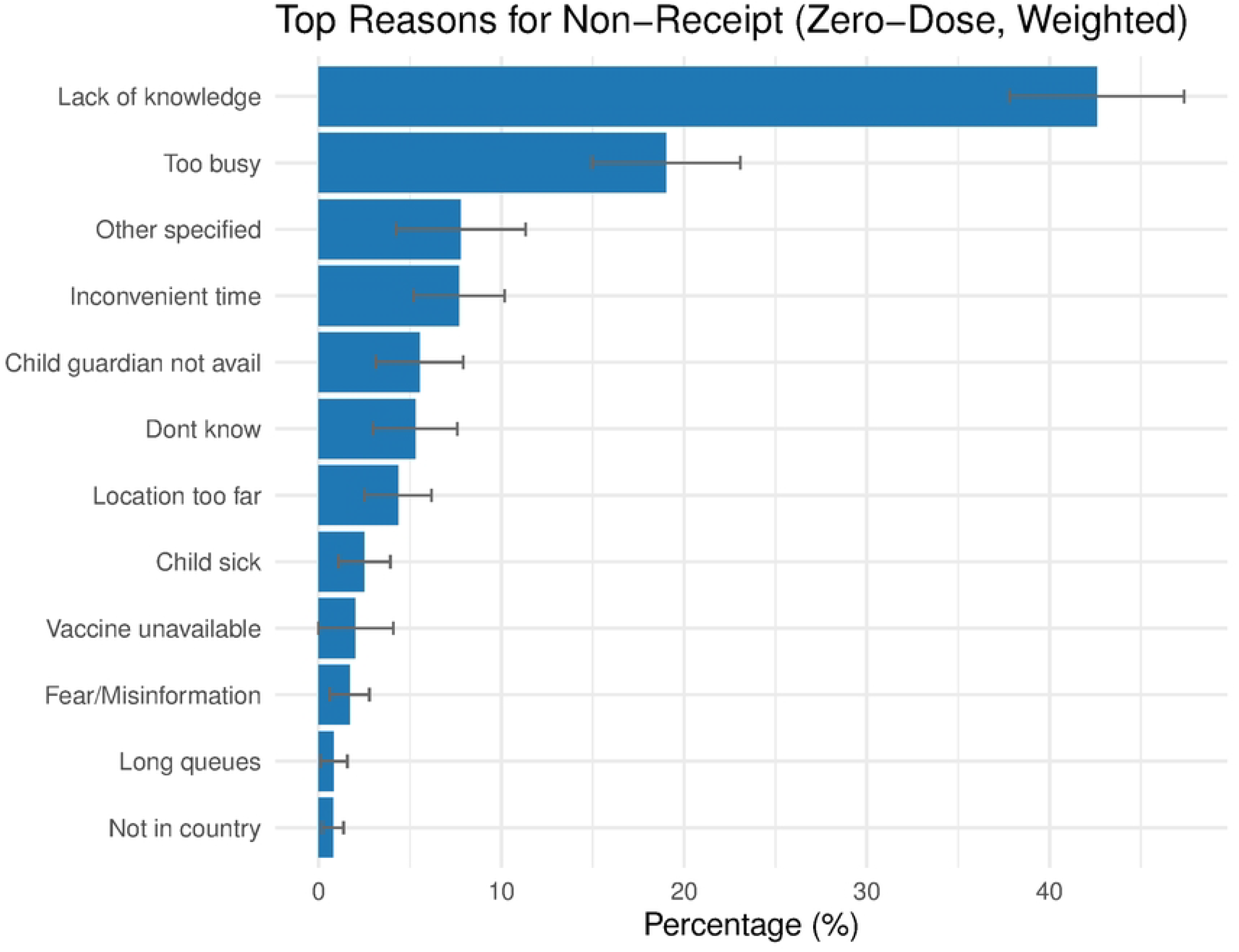
Weighted Reasons for Non-Receipt of Measles-Rubella Vaccine Among Zero-Dose Children, Zambia, 2024 PCCS.

## Discussion

Zambia’s national MR zero-dose prevalence of 11.97%, with high-burden provinces such as Central (19.15%) and Western (17.71%), reflects persistent geographic inequities and structural barriers, including limited access, low campaign awareness, and social vulnerabilities like maternal absence, common in LMICs. This analysis highlights three critical findings: first, clustering of zero-dose children in specific provinces and among older children; second, the dominance of systemic access barriers such as distance and awareness; and third, the overlap of MR zero-dose status with complete lack of other vaccines (88.75%), signalling service delivery failures across the immunisation system rather than isolated campaign shortfalls.

In regional context, Zambia’s 2024 burden (∼41,000 zero-dose children) places it mid-range, higher than countries such as Botswana or Eswatini, but below large-burden nations like Ethiopia and Angola (5). This comparative framing underscores the need for targeted but feasible interventions to reach IA2030 zero-dose reduction goals.

Older children and those with absent or deceased mothers were significantly more likely to be zero-dose, consistent with global evidence linking maternal absence/orphanhood to vaccination gaps (8). These associations may reflect mechanisms such as reduced caregiver decision-making capacity, opportunity costs during agricultural seasons, or limited household support structures. By contrast, socioeconomic status and caregiver education were not significant, suggesting that structural determinants such as distance, service timing, and awareness are stronger drivers than demand-side constraints in this setting.

The co-occurrence of zero-dose MR with absence of other antigens emphasises that Zambia’s challenge lies in routine immunisation system failures. Weak microplanning, poor integration between SIAs and RI, and inconsistent card retention limit opportunities for catch-up. This aligns with global analyses indicating that SIAs often overestimate their independent contribution when RI remains weak (10,11).

Importantly, our interpretation of COVID-19–related disruptions requires clarification. Evidence shows that measles vaccination was strongly protective, with vaccinated children having markedly lower mortality (aOR ∼0.06 compared to unvaccinated (15). Zambia’s pandemic-era gaps therefore exacerbated measles outcomes by reducing coverage, not by conferring protection to unvaccinated children.

Access barriers were central with children living >2 hours from a facility had a prevalence of 26% versus 10% within 15 minutes representing ∼16 percentage-point difference (aOR 3.20). Awareness gaps also remain striking, with 42.6% of caregivers of zero-dose children reporting they had not heard of the campaign. Together, these patterns suggest that reducing travel time and improving health worker–led communication could yield greater gains than socioeconomic-targeted messaging alone.

Generalisability is high for LMIC settings with dispersed rural populations, weak outreach systems, and heavy reliance on SIAs. In dense urban contexts with abundant fixed sites, demand-side determinants (e.g., confidence, complacency) may be more salient, suggesting that Zambia’s lessons apply most strongly to rural, outreach-dependent systems.

This study makes three novel contributions: (i) it quantifies the overlap of MR zero-dose with all-antigen zero-dose nationally, (ii) it demonstrates the primacy of access and awareness barriers after adjusting for stratification, and (iii) it identifies older children and those without maternal caregivers as particularly high-risk groups requiring tailored catch-up strategies.

## Policy Implications

To close gaps, Zambia should:

1. Strengthen proactive communication through health-worker-led, locality-specific campaigns to raise awareness by ≥15 percentage points in high-burden areas.
2. Expand proximity solutions, such as monthly mobile and outreach sessions, to halve >2-hour travel catchments.
3. Integrate SIAs and RI, ensuring all SIA posts conduct defaulter tracing and on-site catch-up for routine antigens, with a target of ≥80% card verification.
4. Adopt geospatial microplanning, updating hotspot maps quarterly to prioritise older children and households lacking maternal caregivers.
5. Deploy digital tools, such as mobile health apps for defaulter tracking and AI-driven hotspot mapping, to enhance microplanning, building on Nigeria’s 20% gap reduction (16).
6. Adapt the cholera control model (17) for measles, using surveillance and community coordination to boost RI in rural hotspots.

By embedding these equity-driven reforms into Zambia’s immunisation strategy, SIAs can serve as true platforms for strengthening RI, closing systemic gaps, and accelerating progress toward measles elimination.

## Limitations

The study has several limitations. Reliance on caregiver recall for 88.3% of vaccination status assessments introduces potential recall bias, which may overestimate or underestimate true prevalence, though this reflects real-world challenges in settings with poor immunization card retention and should be mitigated in future studies with improved card systems. Small sample sizes in some subgroups, particularly for deceased mothers and certain provinces, limit the precision of stratified estimates. Additionally, district-level variations may be masked by provincial-level reporting, potentially obscuring important local patterns that could inform targeted interventions.

## Conclusion

The 2024 Zambia PCCS revealed that nearly 12% of children aged 9–59 months remained MR zero-dose, with the majority excluded from all vaccinations. Disparities were concentrated in rural and high-burden provinces, among older children, and in households lacking maternal caregivers. Structural barriers—including campaign unawareness, long travel times, and seasonal labour conflicts—contributed significantly to non-vaccination.

These findings underscore the need for community-centred, equity-driven strategies that extend beyond campaign-based delivery to strengthen routine immunisation system. By addressing both demand- and supply-side barriers, Zambia can accelerate progress toward measles elimination and contribute to global zero-dose reduction targets under IA2030.

## Data Availability

All data underlying the findings are available upon request

## Acknowledgments

The authors extend their sincere gratitude to the staff of the Zambia Ministry of Health, WHO Zambia Country Office, UNICEF Zambia, and the Zambia Statistics Agency (ZAMSTATS) for their invaluable technical and logistical support throughout the 2024 Post-Campaign Coverage Survey. We also acknowledge the dedication of the data collectors and community health workers whose efforts made this study possible.

## Supplementary Material

**Supplementary Table S1:** Zero-Dose Measles-Rubella Prevalence by Demographic and Geographic Characteristics, Zambia, 2024 PCCS.

| Variable | Prevalence (%) | 95% CI (%) | Odds Ratio (95% CI) |
| --- | --- | --- | --- |
| **Age Group** |  |  |  |
| 0–12 months | 10.50 | 9.32–11.68 | Reference |
| 13–24 months | 12.20 | 10.83–13.57 | — |
| 25–36 months | 13.80 | 12.23–15.37 | — |
| >36 months | 14.99 | 13.25–16.73 | 1.60 (1.27–2.00) |
| **Gender** |  |  |  |
| Male | 11.90 | 10.92–12.88 | — |
| Female | 12.04 | 11.04–13.04 | — |
| **Urban/Rural Status** |  |  |  |
| Rural | 13.02 | 12.04–14.00 | Reference |
| Urban | 10.17 | 9.29–11.05 | 0.76 (0.63–0.92) |
| **Province** |  |  |  |
| Central | 19.15 | 15.84–22.46 | — |
| Western | 17.71 | 15.14–20.28 | — |
| Muchinga | 14.03 | 11.56–16.50 | — |
| Northern | 13.34 | 10.93–15.75 | — |
| North Western | 12.40 | 10.05–14.75 | — |
| Luapula | 11.89 | 9.58–14.20 | — |
| Lusaka | 10.99 | 8.73–13.25 | — |
| Southern | 10.13 | 7.93–12.33 | — |
| Eastern | 9.32 | 7.20–11.44 | — |
| Copperbelt | 6.69 | 4.67–8.71 | — |
| **Maternal Status** |  |  |  |
| Present | 11.00 | 10.06–11.94 | Reference |
| Absent | 19.73 | 15.85–23.61 | 1.74 (1.33–2.27) |
| Deceased | 24.17 | 12.60–35.74 | 2.40 (1.23–4.68) |
| **Household Size** |  |  |  |
| Small (1–4) | 12.29 | 10.72–13.86 | — |
| Medium (5–8) | 11.17 | 9.88–12.46 | — |
| Large (≥9) | 14.18 | 11.63–16.73 | — |

**Note:** Prevalence and 95% confidence intervals (CIs) are weighted estimates from the 2024 Post-Campaign Coverage Survey (PCCS). Odds ratios (ORs) with 95% CIs are shown for significant associations ( $p < 0.05$ ) in survey-weighted logistic regression, with reference categories as indicated. Dashes (—) indicate no OR calculated due to non-significant association ( $p > 0.05$ , e.g., Gender, Household Size) or lack of regression data (e.g., Province). CIs for deceased mothers are wider due to smaller sample sizes.

**Supplementary Table S2:** Zero-Dose Measles-Rubella Prevalence by Socioeconomic and Access Characteristics, Zambia, 2024 PCCS.

| Variable | Prevalence (%) | 95% CI (%) | Odds Ratio (95% CI) |
| --- | --- | --- | --- |
| **Guardian Education** |  |  |  |
| None | 12.22 | 10.89–13.55 | — |
| Primary | 11.33 | 9.82–12.84 | — |
| Secondary | 9.40 | 6.32–12.48 | — |
| Higher | 15.53 | 11.75–19.31 | — |
| **Campaign Awareness** |  |  |  |
| Heard | 10.20 | 9.20–11.20 | Reference |
| Not Heard | 18.50 | 16.17–20.83 | 2.00 (1.65–2.42) |
| **Time to Health Facility** |  |  |  |
| <15 min | 9.59 | 8.00–11.18 | Reference |
| 15–30 min | 9.86 | 8.04–11.68 | — |
| 30–60 min | 12.61 | 10.32–14.90 | 1.35 (1.02–1.79) |
| 1–2 hrs | 14.28 | 12.12–16.44 | 1.55 (1.20–2.00) |
| >2 hrs | 25.71 | 10.94–40.48 | 3.20 (1.43–7.16) |
| **Reasons for non-receipt** |  |  |  |
| Lack of awareness | 42.58 | 37.80–47.36 | — |
| Too busy | 19.02 | 14.96–23.08 | — |
| Inconvenient time | 7.69 | 5.20–10.18 | — |
| **Receipt of Other Vaccines** |  |  |  |
| Zero-Dose (No Other Antigens) | 88.75 | 85.81–91.69 | — |
| Received Other Antigens | 11.25 | 8.31–14.19 | 0.26 (0.08–0.91) |
| **Religion** |  |  |  |
| Catholic | 9.95 | 7.62–12.28 | — |
| Protestant | 12.50 | 11.44–13.56 | — |
| Others | 9.84 | 6.74–12.94 | — |

**Note:** Prevalence and 95% confidence intervals (CIs) are weighted estimates from the 2024 Post-Campaign Coverage Survey (PCCS). Odds ratios (ORs) with 95% CIs are shown for significant associations ( $p < 0.05$ ) in survey-weighted logistic regression, with reference categories as indicated. Dashes (—) indicate no OR calculated due to non-significant association ( $p > 0.05$ , e.g., Guardian Education, Religion, Reasons for Non-Receipt) or lack of regression data. CIs for >2 hrs travel time are wider due to smaller sample sizes. Muslim prevalence (0.00%) was excluded due to unreliable estimates from small sample size. Others in Religion includes African Traditional Religion, Non-Religious, and Other Religious Groups.

